# 6-month aerobic walking training increases T1w/T2w signal in the white matter of healthy older adults

**DOI:** 10.1101/2020.11.09.20228627

**Authors:** Andrea Mendez Colmenares, Michelle W Voss, Jason Fanning, Elizabeth A Salerno, Neha P Gothe, Michael L Thomas, Edward McAuley, Arthur F Kramer, Agnieszka Z Burzynska

**Affiliations:** Department of Psychology/Molecular, Cellular and Integrative Neurosciences, Colorado State University, Fort Collins, CO, USA; Department of Psychological and Brain Sciences, University of Iowa, Iowa City, IA, USA; Department of Health and Exercise Sciences, Wake Forest University, Winston-Salem, NC, USA; Division of Public Health Sciences, Department of Surgery, Washington University School of Medicine in St. Louis, St. Louis, MO, USA; Department of Kinesiology and Community Health, Beckman Institute for Advanced Science and Technology, University of Illinois at Urbana-Champaign, Urbana, IL, USA; Department of Psychology, Northeastern University, Boston, MA, USA; Department of Human Development and Family Studies/Molecular, Cellular and Integrative Neurosciences, Colorado State University, Fort Collins, CO, USA

**Keywords:** aging, white matter, aerobic exercise, plasticity, clinical trial

## Abstract

White matter (WM) deterioration is an important mechanism of cognitive decline in healthy aging and dementia. Engaging in aerobic exercise to improve cardiorespiratory fitness (CRF) is considered one of the most promising ways to improve cognitive and brain health in aging. Yet, no randomized controlled trials have reported benefits of aerobic exercise interventions on WM microstructure measured with diffusion tensor imaging. Here, we studied the effects of a 6-month exercise intervention (clinical trial NCT01472744) on WM of 180 cognitively healthy older adults (60–79 years) using the ratio of calibrated T1-weighted image to T2-weighted images (T1w/T2w). Participants were randomized to one of four groups including a low intensity activity with complex cognitive demands (Dance), Walking, Walking + nutritional supplement or an active control. Results showed that a 6-month aerobic walking and dance intervention produced positive changes in the T1w/T2w with significant time-by-group interactions in the total WM, the genu and splenium of the corpus callosum, forceps minor, cingulum, relative to an active control condition. In contrast, we observed a decline in T1w/T2w in the majority of WM regions in the active control group. Lastly, a positive change in the T1w/T2w in the genu of the corpus callosum correlated with a positive change in episodic memory in the Walking but not in the control group; however, there were no associations between change in the T1w/T2w and change in CRF. Together, our findings suggest that the T1w/T2w may be a sensitive metric to detect short-term within-person changes in the WM and intervention-induced WM plasticity in the adult human brain.

## Introduction

White matter (WM) provides a structural scaffold for brain function and cognition (Burzynska et al., 2011, 2013). The integrity of axons and myelin sheaths determines the speed and synchrony of action potentials, and thus is critical to successful information processing (Chorghay et al., 2018). Cortical “disconnection” (Bartzokis, 2004) due to WM degeneration is widely considered one of the primary mechanisms of age-related cognitive decline (Bartzokis, 2004; Felts et al., 2018). Our recent longitudinal study among healthy older adults showed that declines in WM integrity can be observed in just 6 months (Burzynska et al., 2017). In addition, cross-sectional studies have found that WM deterioration is more pronounced in Alzheimer’s disease (Nasrabady et al., 2018), suggesting a role in the pathophysiology of dementia (Brickman et al., 2015).

Given the well-established evidence of the role of WM degeneration in cognitive aging it is imperative to study lifestyle factors that may prevent or slow down WM decline. A key question is whether WM deterioration is reversible or malleable. Accumulating evidence suggests that WM is more plastic than initially thought (Sampaio-Baptista & Johansen-Berg, 2017). For instance, studies in rodents show experience-related changes in oligodendrocyte differentiation (Simon et al., 2011), myelination (Chorghay et al., 2018), and axonal diameter (Bobinski et al., 2011). In adult humans, engaging in aerobic exercise to improve cardioespiratory fitness (CRF) is considered one of the most promising ways to improve cognitive function (Kramer & Colcombe, 2018), brain functional connectivity (Voss et al., 2016), and reverse age-related brain atrophy (Erickson et al., 2011). Because myelin production and maintenance is metabolically expensive (Rosko et al., 2019), oligodendrocytes are vulnerable to inflammation (Perry & Holmes, 2014), oxidative stress (Giacci & Fitzgerald,2018), and microvascular changes (Tan et al., 2019), all of which are hallmarks of brain aging (Bartzokis, 2011; Nasrabady et al., 2018). However, a recent systematic review among eleven randomized controlled trials reported that aerobic exercise reduced inflammatory markers in older adults (Zheng et al., 2019). Similarly, evidence from a 12-week aerobic exercise intervention (Murrell et al., 2013) and a 7-month aerobic exercise randomized controlled trial (Vicente-Campos et al., 2012) reported improved cerebrovascular reactivity, likely due to improved endothelial function and greater nitric oxide bioavailability.

Surprisingly, randomized controlled trials of both cognitively normal older adults (Burzynska et al., 2017; Clark et al., 2019; Voss et al., 2013) and individuals with mild cognitive impairment or at risk of Alzheimer’s Disease (Fissler et al., 2017; Tarumi et al., 2020; Venkatraman et al., 2020) reported no benefits of 6- to 24-month aerobic exercise interventions on WM microstructure, measured as fractional anisotropy (FA) with diffusion tensor imaging (DTI). A possible explanation for these negative findings is that DTI not only reflects multiple histological processes but it is also affected by the presence of crossing fibers within a voxel (Jeurissen et al., 2013; Jones et al., 2013). Since aging is associated with multiple processes (Liu et al., 2017), such as axonal degeneration, myelin breakdown, glia infiltration, diffusion signal may be insensitive to short-term changes in the adult WM.

A more recent alternative to DTI is the ratio of the standardized T1 and T2-weighted images (T1w/T2w) (Ganzetti et al., 2014; Glasser & van Essen, 2011). In T1-W and T2-W imaging, tissue contrast is created by optimizing the sensitivity of the signal to the differences in the relaxation times of these tissues (Deoni, 2010). Thus, because myelin increases the signal in T1-W images but decreases the signal in T2-W images, the division of the T1-W by the T2-W exploits the shorter relaxation times of myelin and can provide a myelin enhanced contrast (Glasser & van Essen, 2011; Harkins et al., 2016). The T1w/T2w has been shown to be sensitive to the vulnerability of WM in cognitively healthy APOE-4 carriers (Operto et al., 2019) and in neurodegenerative disorders such as multiple systems atrophy (Sugiyama et al., 2020) and multiple sclerosis (Cooper et al., 2019). This suggests that T1w/T2w may be sensitive to functional aspects of WM, offering a new index of WM integrity. Therefore, this study is the first to apply this novel index of WM health to study longitudinal 6-month changes and the effects of aerobic walking or dance intervention on WM microstructure in cognitively normal older adults (clinical trial NCT01472744).

First, given our earlier DTI findings on short-term changes in the aging WM (Burzynska et al., 2017), we hypothetized that T1w/T2w signal would decline or remain stable over 6 months in the control group. Next, we predicted that engaging in a 6-month aerobic walking or dance training (as compared to the strength/stretching/stability control group) would result in lesser decline or an increase in the T1w/T2w signal. We expected that dance would produce a smaller effect size compared to aerobic walking due to its reduced aerobic intensity and effect on CRF than aerobic walking (Voss et al., 2019). Furthermore, we expected that change in CRF—the main variable manipulated by the intervention—would be associated with change in T1w/T2w in the dancing or walking groups. Finally, we expected the change in T1w/T2w to be behaviorally relevant, namely, related to change in cognition.

## Methods

### Participants

Participants were 247 community-dwelling older adults (average age of 65 yrs., 68% women) enrolled in a 24-week randomized controlled exercise trial that examined the effects of aerobic exercise on cognitive performance and brain health. The trial is registered with United States National Institutes of Health ClinicalTrials.gov (ID: <u>NCT01472744</u>). Individuals were eligible to participate if they met the following inclusion criteria: (a) 60–80 years-old; (b) able to read and speak English; (c) scored <10 on the geriatric depression scale (GDS-15); (d) scored ≥ 75% right-handedness on the Edinburgh Handedness Questionnaire; (e) demonstrated normal or corrected-to-normal vision of at least 20/40 and no color blindness; (f) low-active (i.e., participated in 30 or min of moderate physical activity fewer than 2 days per week over the past 6 months); (g) local to the study location for the duration of the program; (h) willing to be randomized to one of four interventions; (i) not involved in another physical activity program; and (j) scored >21 on the Telephone Interview of Cognitive Status questionnaire and >23 on the Mini Mental State Exam (Fong et al., 2009). Eligibility also included meeting inclusion criteria for completing a magnetic resonance imaging (MRI) assessment, consisting of: (a) free from neurological disorders; (b) no history of stroke, transient ischemic attach, head trauma or surgeries including the removal of brain tissue; and (c) no implanted devices or metallic bodies above the waist. Thus, our sample consisted of healthy, community-dwelling, typically low active older adults. For more information on participant recruitment and screening, see (Baniqued et al., 2018; Burzynska et al., 2017; Ehlers et al., 2016, 2017; Fanning et al., 2016; Voss et al., 2018). Participants underwent a series of MRI imaging, cognitive, and cardiorespiratory testing, before and after the 6-month intervention program.

The study was approved by and carried out in accordance with the recommendations of the Institutional Review Board at the University of Illinois at Urbana-Champaign with written informed consent from all participants. All participants provided written informed consent in accordance with the Declaration of Helsinki.

### Intervention

After all baseline data were collected, participants were assigned to one of four interventions implemented over four waves from October 2011 to November 2014. Participants were randomized using a computer data management system and baseline-adaptive randomization scheme, taking into account equal distributions of age and gender (Begg & Iglewicz, 1980). Participants in all conditions attended three 1-h exercise sessions per week for 24 weeks (∼6 months)(Burzynska et al., 2017; Ehlers et al., 2017). The four intervention groups included: **Strength/stretching/stability (SSS)** involved exercises designed to improve flexibility, strength, and balance with the aid of yoga mats and blocks, chairs, and resistance bands, specifically designed for individuals 60 years of age and older. This intervention served as the active control group to account for the social engagement and novelty in the other interventions, with the difference that SSS was not aimed to increase CRF. The **Walking** intervention was designed to increase CRF. Thus, it involved walking sessions at 50–60% of maximal heart rate, as ascertained from a maximal graded exercise test. Walking duration increased from 20 to 40 min during the first 6 weeks of the program. During the remaining 18 weeks, participants walked for 40 min at 60–75% of their maximal heart rate each session. Frequent assessment of heart rate, using either palpation or Polar Heart Rate Monitors, and rating of perceived exertion ensured that participants’ exercise was performed at the prescribed intensity. Exercise logs were completed after each exercise session. The **Walking + Nutrition** group, in addition to the above walking intervention, received a nutritional supplement containing antioxidants, anti-inflammatories, vitamins, minerals, and beta alanine (Abbott Nutrition, Abbott Park, Illinois). Beta-alanine is thought to promote an increase in lean muscle mass (Zoeller et al., 2007), thereby enhancing the effect of increased CRF. However, the analyses of the primary outcomes indicated no differences in gain in CRF between the walking interventions (Baniqued et al., 2018; Ehlers et al., 2017; Voss et al., 2019) therefore, Walking and Walking + Nutrition were collapsed for the present analyses. The **Dance** intervention was designed to provide simultaneous cognitive and social enrichment combined with aerobic physical activity (PA). The choreographed dance combinations became progressively more challenging over the course of the 6-months program. Group social dance styles were selected (i.e., Contra and English Country dancing) to minimize lead-follow roles. In each session, participants learned ∼4 dances and recorded their heart rate and perceived exertion after each dance. Each participant learned and alternated between two roles for each dance, increasing the cognitive challenge.

### Cardiovascular variables

Cardiorespiratory fitness (CRF) was assessed before and after the intervention on a motor-driven treadmill by employing a modified Balke protocol (graded exercise test). The protocol involved walking at a self-selected pace with incremental grades of 2-3% every 2 minutes. We continuously collected measurements of oxygen uptake, heart rate and blood pressure. We measured oxygen uptake (VO_2_) from expired air samples taken at 30-second intervals until a peak VO_2_ (the highest VO_2_) was attained; test termination was determined by symptom limitation, volitional exhaustion, and/or attainment of VO_2_ peak as established by the American College of Sports Medicine guidelines (American College of Sports Medicine, 2013).

### MRI Acquisition

We acquired images on a 3T Siemens Trio Tim system with 45 mT/m gradients and 200 T/m/sec slew rates (Siemens, Erlangen, Germany). T1-weighted images were acquired using a 3D MPRAGE (TR = 1900 ms; TE = 2.32 ms; TI: 900 ms; matrix = 256 × 256; FOV = 230 mm; 192 slices; 0.9 × 0.9 × 0.9 mm^3^ voxels size; GRAPPA acceleration factor 2). The non-diffusion weighted images from the diffusion-weighted acquisition were used as T2-weighted images (b-value = 0 s/mm^2^, TR = 5500 ms; TE = 98 ms, matrix = 128 × 128; 1.7 × 1.7 x 3 mm^3^ voxels size; GRAPPA acceleration factor 2), because the study protocol did not include a T2-W image scan besides FLAIR, which is suboptimal for the T1w/T2w calculation since it has a decreased grey-WM contrast due to the inversion pulse (Ganzetti et al., 2014). Out of 213 participants who completed the intervention, 180 had good quality MRI data at pre and post-intervention (see Supplementary Material 1 for participant flow for the current analyses). AMC and AZB independently checked for image quality. Images were excluded from the analyses if they had motion or ghost artifacts that affected the grey-WM boundary or image co-registration; 8 subjects were excluded due to brain anatomical concerns that affected image co-registration and could lead to partial volume effects (e.g., ventriculomegaly or asymmetrical ventricles); 8 subjects were excluded due to insufficient brain coverage of their T2-W images for intensity calibration with the MRTool.

#### T1w/T2w calculation

We calculated T1w/T2w images with the MRTool registration-segmentation framework in SPM12 (Wellcome Trust Centre for Neuroimaging, London, UK; (Ganzetti et al., 2014). First, the images were bias corrected to normalize the sensitivity profiles across subjects and scan sessions. The bias correction algorithm included smoothing (60mm) of the intensity inhomogeneity and regularization (10^−4^) to sharpen intensity transitions between image structures. Next, the images were calibrated to standardize their intensity scales across sessions and participants (Ganzetti et al., 2014). We could not use the recommended external calibration (using the eye and temporal muscle) due to insufficient head coverage of the T2-W images. Thus, we used a process known as histogram equalization to rescale the image on the basis of the whole brain intensity distribution (Ganzetti et al., 2014; Glasser & van Essen, 2011). Then, T2-W images in the individual space were co-registered to T1-W images through a 6 degrees of freedom rigid-body transformation. Finally, the T1w/T2w were calculated in individual space using the bias corrected and calibrated images, and then transformed to Montreal Neurological Institute (MNI) space 1mm^3^ in SPM (Ganzetti et al., 2014).

#### Skeletonization and region selection

We used Tract-Based Spatial Statistics (TBSS) in FSL (Smith et al., 2006) to restrict the analyses to the center of WM tracks. This was to minimize the effects of possible partial volume due to individual and age differences in anatomy, to focus the analyses on the normal-appearing WM, and to allow direct comparison with our earlier DTI findings from this sample (Burzynska et al., 2017). We used the non-FA TBSS pipeline for the T1w/T2w images to project them onto the group WM skeleton with a threshold of FA > 0.2, as we described earlier (Burzynska et al., 2017). To confirm that the T1w/T2w voxels were correctly projected onto the WM skeleton, we de-projected all skeletonized T1w/T2w images for visual inspection in subject’s native space; the deprojection was accurate for all participants and regions except for regions 3 and 4 of the corpus callosum in 5 participants, which were treated as missing values.

We extracted T1w/T2w regional values for statistical analyses. Total WM was defined as all voxels on the WM skeleton. We examined the five subsections of the corpus callosum (CC) (Hofer & Frahm, 2006) given the anterior-to-posterior gradient of CC’s vulnerability to aging (Head et al., 2004). Other WM regions included the association fibers connecting regions known to be affected by aging: the fornix (FX), the superior longitudinal fasciculus (SLF), the external capsule (EC), the cingulum (CING), and the uncinate fasciculus (UNC). In addition, we included two other major WM landmarks: the forceps minor (fMIN) and forceps major (fMAJ), containing callosal fibers and thalamic projections to the frontal lobes and the occipital lobes, respectively. The corticospinal tract (CST) represented long projection fibers, typically more resilient to aging but possibly involved in the physical activity interventions. To define fMIN, fMAJ, UNC, SLF and CST on the WM skeleton, we used the tract probability maps from the John Hopkins University WM tractography atlas (Hua et al., 2008); http://cmrm.med.jhmi.edu). We thresholded the tract probability maps at 10-15%, depending on a tract, with the aim to maximize the overlap with WM skeleton but avoid including voxels from neighboring tracts (Figure 1). For the FX and EC, we used the John Hopkins University WM labels in FSL. Finally, since the prefrontal cortex is vulnerable to aging (Head et al., 2004) and its volume and function has been shown to benefit from greater CRF or aerobic exercise (Colcombe & Kramer, 2003; Voss et al., 2013), we defined prefrontal WM using a cutoff of y > 12 in MNI space (Burzynska et al., 2013). Then, we winsorized T1w/T2w values for each region at one percent of their distributions, separately for pre and post-intervention.

**Figure 1.**
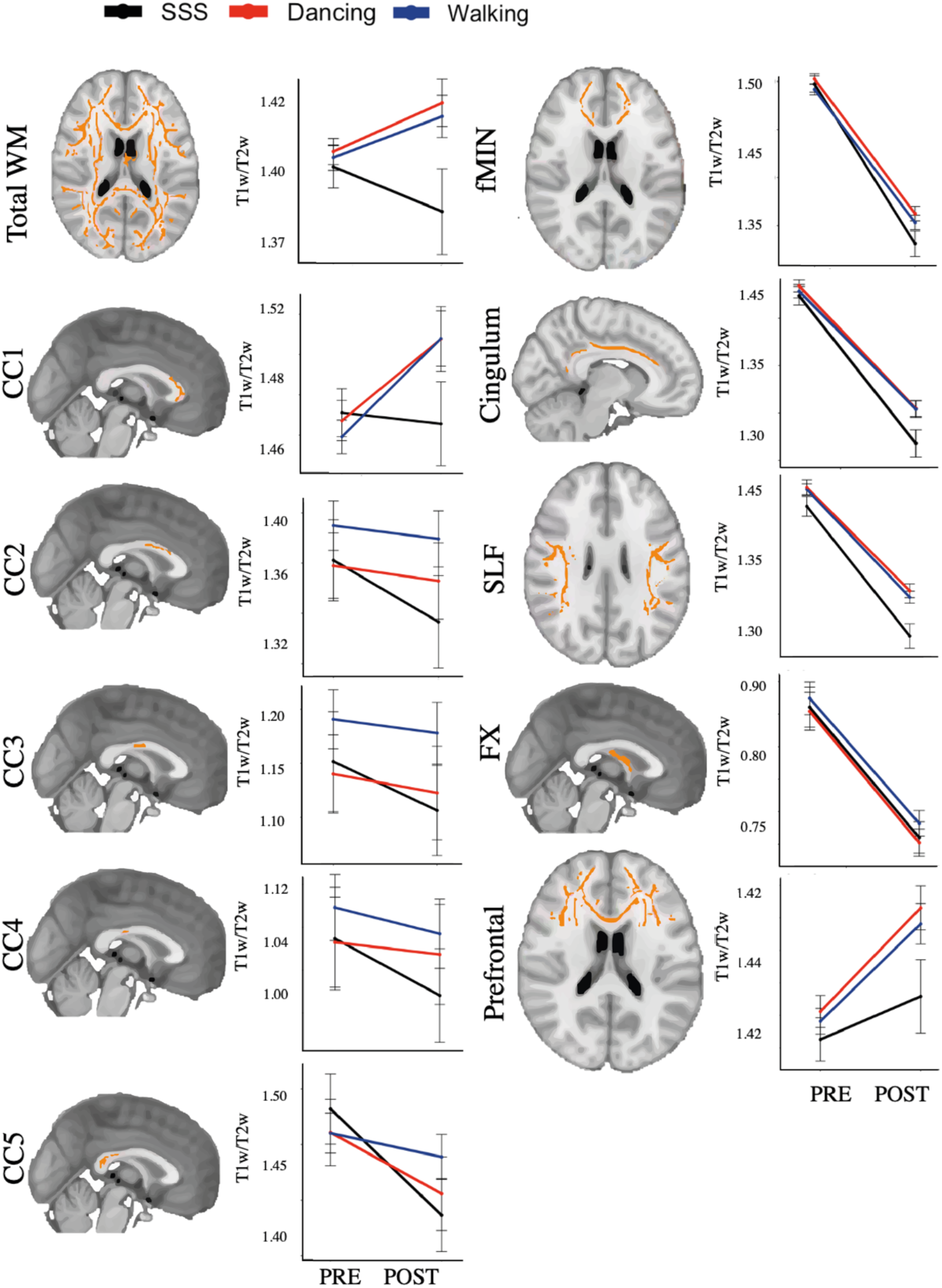
6-month change in T1w/T2w signal in the SSS, Walking and Dance groups *Note*. The points represent the group means at both preintervention (PRE) and postintervention (POST) for each intervention group, and error bars represent 95% confidence intervals.

Finally, we used Shapiro-Wilk tests with visual inspection of the histograms and QQ plots to assess the normality and distribution of the T1w/T2w data. Frequency histograms of T1w/T2w in the whole sample showed a bimodal distribution in the following regions: forceps major, uncinate fasciculus, external capsule and corticospinal tract. This could have diluted the between-groups mean differences in these regions, leading to underestimation of the intervention effects and overestimation of the effects of time. Therefore, we removed these four regions from the main analyses and included the analyses in the Supplementary Material 2.

### Cognitive assessment

Cognitive assessment included the Virginia Cognitive Aging (VCAP) battery (Salthouse, 2009, 2017) and two additional experimental executive function tasks (task switching and spatial working memory (Baniqued et al., 2018; Erickson et al., 2011). We have previously shown that the task switching and spatial working memory load on the reasoning construct of the VCAP battery (Baniqued et al., 2018; Voss et al., 2018). Thus, we grouped VCAP reasoning tasks (matrix reasoning, Shipley abstraction, letter sets, spatial relations, paper folding, and form boards) with the task switching and the spatial working memory task to create an executive function composite (Baniqued et al., 2018; Voss et al., 2018). In addition to the executive function composite, the VCAP assessed episodic memory (word recall, paired associates, logical memory tasks), perceptual speed (digit symbol substitution, letter comparison, pattern comparison), and vocabulary (Wechsler Adult Intelligence Vocabulary, picture vocabulary, and synonym/antonym). We used the vocabulary construct only for sample description, because there is no evidence linking physical activity interventions with gains in crystallized abilities.

We winsorized values for each task at one percent of their distributions, separately for pre and post-intervention. Then, we expressed both pre and post-intervention individual values as standardized scores (z-scores) using the mean and standard deviation of the pre-intervention distribution. Finally, we calculated composite scores for both pre- and post-intervention as mean z-scores of tasks within each cognitive domain (using the mean and standard deviation from the pre-assessment).

### Statistical analyses

We combined the two Walking intervention groups because earlier results indicated no difference in CRF gains between the Walking and Walking + Nutrition groups (Voss et al., 2018). We used linear mixed-effects (LME) models with parameter estimates fitted using the R lme4 package (Bates et al., 2015) to compare change in T1w/T2w between the three groups (Walking, Dance, and SSS). Models included fixed effects of time, group, and the time-by-group interaction as well as random intercepts. The group factor was coded using Helmert contrasts. This allowed us to compare the SSS control against the average of all the Walking and Dance groups. Then, to contrast the effects of Walking vs SSS and Dance vs. SSS we fitted additional LME models using a contrast matrix with dummy codes for the three groups, such that SSS was the reference. We standardized all quantitative variables, but not factors, to create partially standardized regression coefficients. The standardization of our variables rendered regression coefficients (*β*) that are loosely interpreted like correlation coefficients in terms of effect size (Ferguson, 2009). We tested the assumptions of the LME models by visually inspecting the normality of residuals, as well as the distribution of the residuals vs. fitted values.

For correlational analyses, 6-month change scores in the variables of interest were calculated as the post-intervention z-score minus pre-intervention z-score (note that we used the pre mean and standard deviation to transform both pre and post data). We used partial Pearson’s correlations in R ppcor to study the associations between change in T1w/T2w and cognition (controlling for age, sex and education), and between change in T1w/T2w and CRF (controlling for age and sex) within each intervention group. Because these correlational analyses were exploratory, we corrected for multiple comparisons using the false discovery rate method as implemented by p.adjust (p.value, method=“fdr”) in R. Statistical significance was accepted at p<0.05 for two-tailed tests. For the size of our correlations and effects, we assumed 0.50 as a large-sized effect, 0.30 a medium-sized effect and 0.10 a small-sized effect (Cohen, 1992).

We created figures using the ggplot function in the ggplot2 package (Wickham, 2009) and the multiplot function within the coefplot package (Lander, 2016). All statistical analyses were completed using R version 4.0.1.

## Results

One-way ANOVA showed no baseline differences in age, gender, education, cardiorespiratory fitness, and regional T1w/T2w values between the three groups (Supplementary Material 3), indicating successful randomization.

### Intervention effects

Comparing the combined Walking and Dance groups vs. SSS, we found a significant time-by-intervention interaction in 6 out of the 11 WM regions, which included: total WM, the anterior and posterior subsections of the corpus callosum, forceps minor, cingulum and superior longitudinal fasciculus (Table 1). A comparison between Walking and SSS showed significant time-by-intervention interactions in the total WM, genu and splenium of the corpus callosum, forceps minor and the cingulum. Similarly, the models comparing Dance and SSS showed significant time-by-intervention interactions in the total WM and the genu of the corpus callosum, and at trend level in the cingulum. Using Helmert contrasts we compared the effect sizes quantifying the intervention effects between the Dance vs. Walking groups and we did not find significant differences between groups (Supplementary Material 4). Together, our results showed positive intervention-related changes in the T1w/T2w when compared to the SSS control (Figure 1), with larger effect sizes in the Walking group.

**Table 1.** Time-by-group interactions in WM T1w/T2w

| Region | Walking + Dance vs. SSS |  |  | Walking vs. SSS |  |  | Dance vs. SSS |  |  |
| --- | --- | --- | --- | --- | --- | --- | --- | --- | --- |
| | $\beta$ | SE | $p$ | $\beta$ | SE | $p$ | $\beta$ | SE | $p$ |
| Total | 0.26 | 0.11 | **0.02** | 0.25 | 0.11 | **0.03** | 0.27 | 0.13 | **0.04** |
| CC1 | 0.24 | 0.09 | **0.01** | 0.22 | 0.10 | **0.02** | 0.22 | 0.11 | **0.05** |
| CC2 | 0.09 | 0.06 | 0.12 | 0.09 | 0.06 | 0.13 | 0.09 | 0.06 | 0.19 |
| CC3 | 0.05 | 0.05 | 0.26 | 0.06 | 0.05 | 0.26 | 0.05 | 0.06 | 0.38 |
| CC4 | 0.06 | 0.04 | 0.13 | 0.05 | 0.04 | 0.25 | 0.07 | 0.05 | 0.25 |
| CC5 | 0.14 | 0.06 | **0.01** | 0.18 | 0.06 | **0.01** | 0.10 | 0.07 | 0.12 |
| Prefrontal | 0.17 | 0.10 | 0.10 | 0.16 | 0.10 | 0.14 | 0.18 | 0.12 | 0.14 |
| fMIN | 0.14 | 0.07 | **0.03** | 0.15 | 0.07 | **0.04** | 0.14 | 0.07 | 0.20 |
| CING | 0.15 | 0.06 | **0.02** | 0.16 | 0.07 | **0.02** | 0.14 | 0.02 | 0.07 |
| SLF | 0.15 | 0.07 | **0.05** | 0.13 | 0.07 | 0.09 | 0.16 | 0.09 | 0.08 |
| FX | 0.01 | 0.05 | 0.86 | 0.01 | 0.05 | 0.70 | -0.03 | 0.05 | 0.94 |

*Note.* SE= standard errors, CC= corpus callosum, fMIN= forceps minor, CING= cingulum, SLF= superior longitudinal fasciculus, FX= fornix. $\beta$ are standardized. Bold highlights $p < .05$ . WM regions are explained and visualized in Figure 1.

### 6-month longitudinal change in T1/T2 in the control group

As predicted, we observed a consistent pattern of decline or stability of the T1w/T2w signal over a period of 6 months in the SSS control group in the majority of WM regions. The largest effect sizes were observed in WM regions where we found significant time-by-group interactions (forceps minor and cingulum). Figure 1 shows the means for the T1w/T2w at preintervention and postintervention for each intervention group, while Figure 2 shows the standardized *β*-coefficients for all WM regions for the effects of time in the SSS control. Together, Figures 1 and 2 show a significant negative effect of time on the T1w/T2w in all WM regions except the genu of the corpus callosum and prefrontal WM, which showed no 6-month change.

**Figure 2.**
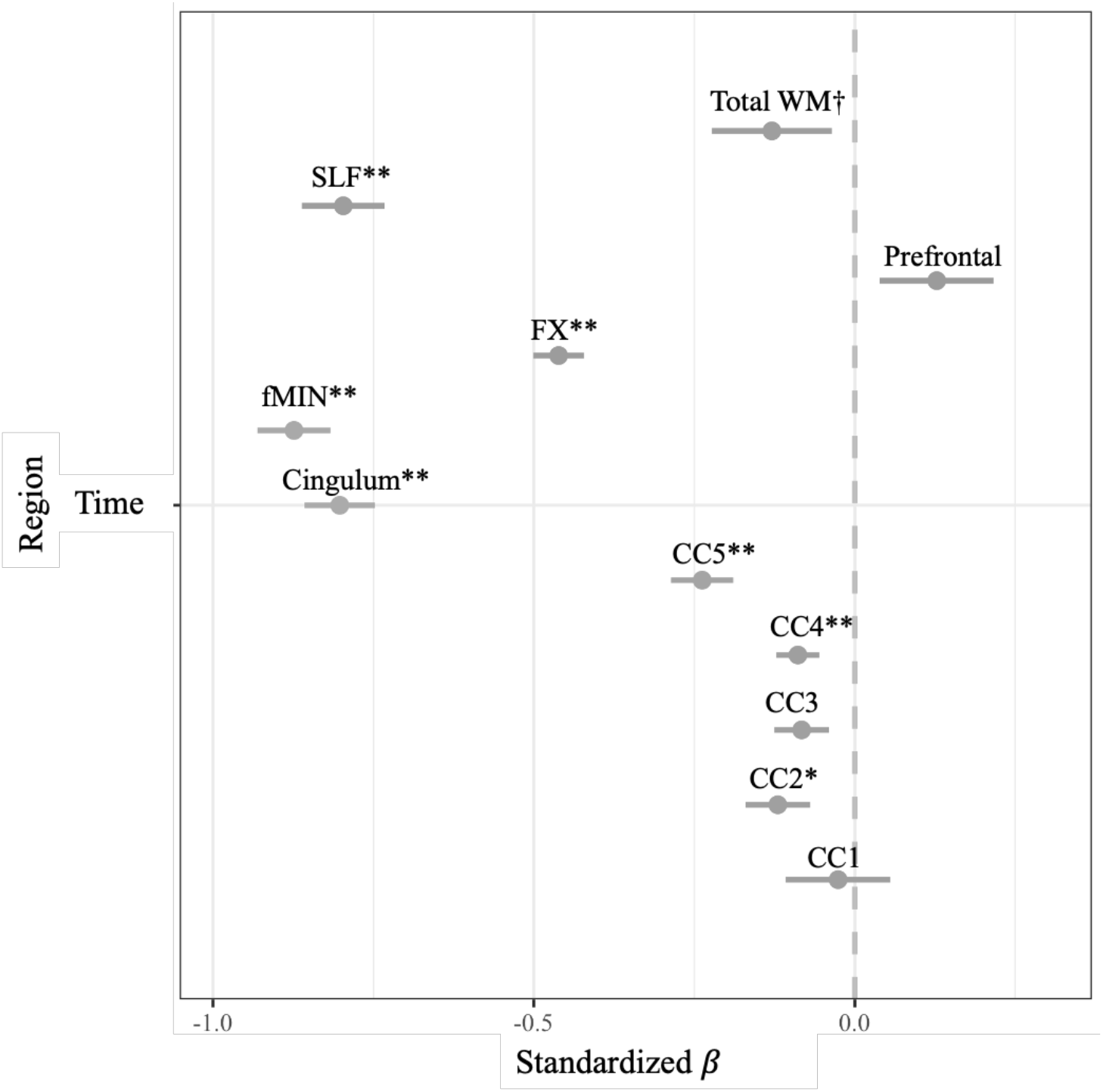
Standardized β coefficients for the fixed effects of time in the SSS control. *Note*. Asterisks indicate †<0.10, \**p*<0.05, \*\**p*<0.01. Error bars represent 95% confidence intervals.

### Change in T1w/T2w and cognition

Because the Walking group produced small to medium sized effects in five WM regions in the LME models, we correlated 6-month change in T1w/T2w in these regions with change in memory, perceptual speed and executive function, while controlling for age, sex, and education. We found that a positive change in T1w/T2w correlated with a positive change in episodic memory, in the genu of the corpus callosum and cingulum, only in the Walking group (Table 2). None of these effects were significant in the SSS group and Dance group after controlling for multiple tests. Lastly, we did not find significant associations between baseline T1w/T2w and baseline cognitive scores (Supplementary Material 5).

**Table 2.** Association between change in T1w/T2w and change in cognitive scores

| Region | Episodic Memory |  |  | Perceptual Speed |  |  | Executive Function |  |  |
| --- | --- | --- | --- | --- | --- | --- | --- | --- | --- |
|  | SSS | Walking | Dance | SSS | Walking | Dance | SSS | Walking | Dance |
|  | n=43 | n=86 | n=51 | n=43 | n=86 | n=51 | n=43 | n=86 | n=51 |
| Total | 0.04 | 0.28* | -0.04 | -0.27 | -0.06 | 0.12 | -0.34 | 0.01 | -0.04 |
| CC1 | -0.12 | 0.27* | -0.04 | -0.21 | 0.10 | 0.09 | 0.10 | 0.10 | 0.10 |
| CC5 | -0.25 | 0.16 | -0.20 | -0.04 | 0.17 | 0.03 | 0.17 | 0.17 | 0.17 |
| fMIN | 0.01 | 0.21 | 0.06 | -0.27 | 0.01 | 0.08 | 0.01 | 0.01 | 0.01 |
| Cingulum | -0.09 | 0.21 | 0.01 | -0.24 | -0.07 | 0.10 | -0.07 | -0.07 | -0.07 |

*Note.* \* $p < 0.05$ , \*\* $p < 0.01$ . CC= corpus callosum, fMIN= forceps minor, CING= cingulum. Partial correlations between change in T1w/T2w and cognition within each intervention group, controlling for age, sex, and education. Significance corrected for FDR.

### T1w/T2w and CRF

Because CRF was the primary physiological outcome of the intervention, we tested whether the intervention-related changes in T1w/T2w were associated with increases in CRF. Pearson partial correlations, controlling for age and sex, revealed no significant associations between change in T1w/T2w and CRF after controlling for multiple tests (Table 3).

**Table 3.**
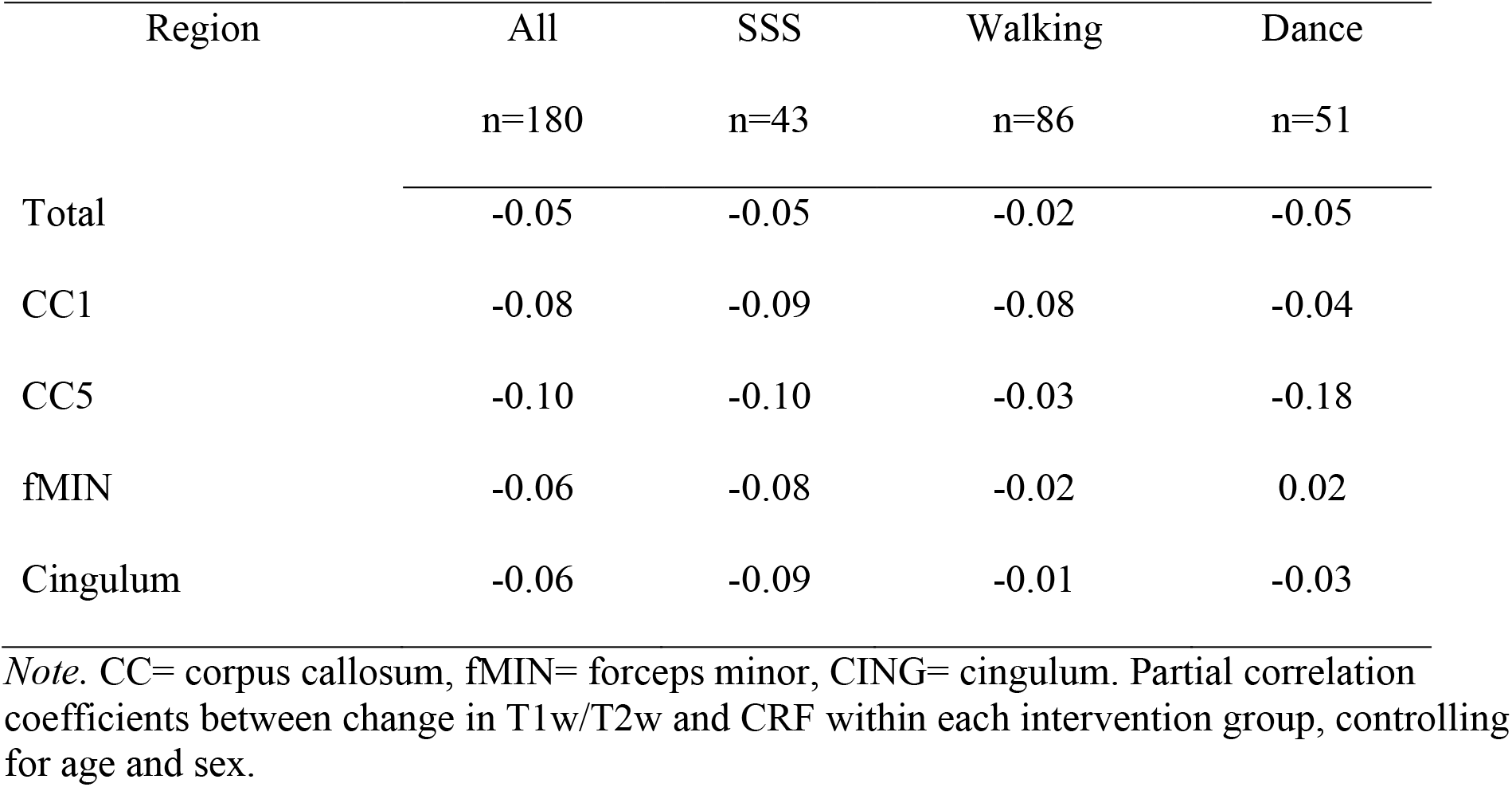
Partial correlation coefficients between change in T1w/T2w and change in CRF

| Region | All | SSS | Walking | Dance |
| --- | --- | --- | --- | --- |
|  | n=180 | n=43 | n=86 | n=51 |
| Total | -0.05 | -0.05 | -0.02 | -0.05 |
| CC1 | -0.08 | -0.09 | -0.08 | -0.04 |
| CC5 | -0.10 | -0.10 | -0.03 | -0.18 |
| fMIN | -0.06 | -0.08 | -0.02 | 0.02 |
| Cingulum | -0.06 | -0.09 | -0.01 | -0.03 |

*Note.* CC= corpus callosum, fMIN= forceps minor, CING= cingulum. Partial correlation coefficients between change in T1w/T2w and CRF within each intervention group, controlling for age and sex.

## Discussion

The present study is the first to use the standardized T1w/T2w to assess adult WM plasticity in a randomized controlled trial. The key findings are:

1. After 6 months, the T1w/T2w showed significant differences between the SSS control and the intervention groups (Dance and Walking) in 6 out of 11 studied WM regions, predominantly among frontal WM regions.
2. The T1w/T2w showed a significant effect of time in the SSS control in 7 WM regions, indicating a spatially wide-spread 6-month decrease in T1w/T2w signal.
3. In the total WM and the genu corpus callosum there was a positive association between change in T1w/T2w and change in episodic memory in the Walking group, but not in the SSS control or the Dance group.
4. The T1w/T2w was not associated with change in CRF when controlling for age and sex.

### Aerobic exercise training increased T1w/T2w in the adult WM

Our results confirmed our hypothesis that the aerobic exercise intervention (Walking and Dance training) resulted in an increase in the T1w/T2w signal, relative to an active control condition. Our findings are in alignment with the evidence supporting a positive relationship between aerobic exercise and brain structure and function (Colcombe et al., 2006; Erickson et al., 2011; Kleemeyer et al., 2016; Voss et al., 2010). However, previous randomized controlled trials using DTI reported no group effects of aerobic exercise (Burzynska et al., 2017; Clark et al., 2019; Voss et al., 2013). To the best of our knowledge, this is the first study to report exercise-induced changes in WM microstructure in healthy older adults.

Interestingly, the effects of the interventions on the T1w/T2w were region-specific. In the Walking group, the T1w/T2w showed positive changes in total WM, the genu and splenium of the corpus callosum, as well as the forceps minor and the cingulum. The majority of these WM regions contain late-myelinating association fibers that interconnect with the prefrontal cortex, with myelination extending into young adulthood (Knyazeva, 2013; Lebel et al., 2019; Pfefferbaum et al., 2000). Because WM regions that myelinate later in development deteriorate earlier with age (Bartzokis et al., 2010; Brickman et al., 2012), our findings suggest that regions vulnerable to aging retain some level of plasticity that can be induced by exercise interventions. Our results are comparable to correlational DTI studies in healthy older adults that showed that aerobic fitness was positively correlated with FA in the body and genu of the corpus callosum (Johnson et al., 2012), and the cingulum bundle (Marks et al., 2011). This study shows promising evidence of neural plasticity of structural connections within the vulnerable frontal and prefrontal regions.

### Dance vs. Walking interventions

Although we observed no difference between change in T1w/T2w between the Walking and Dance group, we observed larger effect sizes in the Walking group for the time-by-intervention interactions. It is tempting to conclude that aerobic walking may be more effective in increasing WM’s T1w/T2w than dancing. However, because the Walking group (n=86) had a larger sample size than the Dance group (n =51), the lack of significant effects in some of the regions could be due to lower statistical power in the models comparing Dance vs. SSS. In addition, the Dance intervention included a large amount of low-intensity instructional time and less intensive aerobic activity. This is consistent with previously observed lower gains in CRF in Dance vs. Walking groups (Voss et al., 2018).

It is important to note that our previous DTI analyses showed that 6-month Dance intervention (but not Walking) resulted in an increase in FA in the fornix, which declined in the corntrol group (Burzynska et al., 2017). It is possible that the Dance intervention elicits changes in WM that are better measured with DTI in the fornix, which is a thin tract containing small-diameter fibers with many unmyelinated axons, and it is surrounded by the lateral ventricles. It is possible that the T1w/T2w cannot capture changes in the fornix because of the partial volume effects induced by the cerebrospinal fluid or because of the microstructural characteristics in this region.

### Within-person changes in WM measured with T1w/T2w

T1w/T2w decreased over a 6-month period in the majority of WM regions in the SSS control, with the largest effect sizes observed in the forceps minor, cingulum and superior longitudinal fasciculus and the smallest effect sizes observed in the anterior corpus callosum. Our results using T1w/T2w are consistent with our earlier findings using DTI. Namely, in a similar set of participants we observed 6-month decline in FA in the in association fibers (e.g., cingulum), limbic system structures (e.g., fornix), and in commisural fibers (corpus callosum) (Burzynska et al., 2017). However, for other regions, our current results suggest a different pattern of age-related decline in the T1w/T2w signal. For instance, we did not find 6-month change in the genu of the corpus callosum, but reported a change in the body and splenium of the corpus callosum. Perhaps, FA is more sensitive to changes in regions with smaller axons that are coherently oriented and densely packed (e.g., genu) (Burzynska et al., 2010, 2017). On the other hand, the T1w/T2w may be better suited to detect longitudinal changes in regions characterized by larger axonal diameter, such as the splenium of the corpus callosum (Lamantia & Rakic, 1990) or by high crossing fibers such as the cingulum bundle or the superior longitudinal fasciculus (Glenn et al., 2016). In sum, our results suggest that the degree of decline in the T1w/T2w varies by region, so it is possible that these regional differences in susceptibility to aging reflect distinct mechanisms of WM aging. However, a systematic examination of this issue using more advanced WM techniques is required to confirm such a speculation.

### Episodic Memory and T1w/T2w

In two out of the five regions with significant intervention effects, we found a positive association between change in T1w/T2w and change in episodic memory in the Walking group. One of these regions was the genu of the corpus callosum, which is known to mediate interhemispheric integration in episodic memory processes. Specifically, the genu of the corpus callosum has been shown to be implicated in the recruitment of the ventrolateral prefrontal cortex and to mediate the relationship between perceptual speed and episodic retrieval in healthy older adults (Bucur et al., 2008). We also observed significant associations in the total WM, possibly driven by the effects observed in all WM regions combined, and in the cingulum (this latter at a trend level). This is consistent with evidence showing that decreased FA in the dorsal cingulum has been associated with episodic memory impairment (Lockhart et al., 2012).

Given the effects of exercise on executive functions and processing speed (Colcombe & Kramer, 2003; Kramer & Colcombe, 2018) and the reliance of processing speed on WM integrity (Chopra et al., 2018), we were surprised to find no associations between change in T1w/T2w and change in these two cognitive domains. Although we observed performance decline trends in the control group for the executive function and processing speed components, these were not statistically significant. Similarly, in our previous work, we did not find significant time-by-group interactions showing training-related changes on the cognitive level (Voss et al., 2018). A systemic meta-analysis confirmed no beneficial effects of short-term aerobic training interventions on these cognitive domains (Young et al., 2015). Overall, our results suggest that the T1w/T2w is capable of detecting positive changes of WM in regions known to be involved in episodic memory processes. However, different modifiers can influence the relationship between aerobic exercise and cognition, such as physical activity intensity, intervention duration, diet, social engagement and genetic factors. Thus, future studies should test the associations between change in T1w/T2w and cognition over longer periods of time while testing for important individual differences that can moderate the effects of aerobic exercise on brain health.

### Cardiorespiratory fitness and WM plasticity

We did not find an association between change in T1w/T2w and change in CRF. Even though numerous studies have reported positive associations between increased CRF and brain function and structure (Kramer & Colcombe, 2018), our results do not support our prediction that increased T1w/T2w signal would be associated with increased CRF, suggesting that the increased T1w/T2w in the aerobic exercise groups is mediated by other mechanisms other than CRF. However, it is important to note that increased CRF is associated with moderate-to-high intensity aerobic exercise, as corroborated by results from this randomized controlled trial showing that the Walking groups increased CRF (Voss et al., 2018). Aerobic exercise training is associated with physiological adaptations that result in increased oxygen-carrying capacity and extraction by tissues, which rapidly improves CRF in the first 5-11 weeks of training (Lundby et al., 2017). Subsequently, smaller improvements in CRF would be expected after months of training (3-12 months) (Erickson et al., 2011; Vidoni et al., 2015; Voss et al., 2018). Because our study sample was composed of healthy individuals, with relatively normal blood pressure values (Supplementary Material 3), it is possible that the associations between CRF and arterial health are stronger among individuals with hypertension or other chronic diseases (Haapala et al., 2020) and thus not observable in our healthy sample. Since vascular health is likely a key mechanism underlying the protective effects of aerobic exercise training on brain health, future studies are needed to further characterize individual differences that mediate or moderate the protective effects of aerobic exercise on the aging WM.

However, we found that increased T1w/T2w correlated with episodic memory in frontal and prefrontal WM regions only in the Walking group. This resembles findings by Ruscheweyh and colleagues, where they found that a 6-month aerobic exercise intervention correlated with increased gray matter volumes (mostly in the prefrontal and cingulate cortex) and episodic memory performance (word list learning), independently of CRF (Ruscheweyh et al., 2011). Our results suggest that any form of aerobic exercise is associated with positive changes in T1w/T2w, but only Walking is associated with better episodic memory functions.

### T1w/T2w as an index of WM health

Because this is the first application using T1w/T2w to assess the effects of a clinical trial, the meaning of our findings need to be interpreted with caution. Although 3–4 weeks of aerobic exercise training was shown to stimulate myelination and increase axonal diameter in mice (Bobinski et al., 2011; Chen et al., 2019), relating changes in T1w/T2w to any particular microstructural mechanism, such as myelin content, is premature. Overall, evidence suggests that the T1w/T2w offers a promising measure of the functional aspects of WM microstructure (Uddin et al., 2019) that shows high concurrent validity after calibration (Arshad et al., 2017). Even though the T1w/T2w was first designed to map myelin content in the human neocortex (Glasser & van Essen, 2011), several biophysical processes in WM contribute to changes in T1w/T2w intensity (Deoni, 2010). The contrast of different tissues in T1-WI and T2-WI arise largely from distinct T1 and T2 relaxation properties of brain tissues (Sharma & Lagopoulos, 2010). However, T1 and T2 relaxations are not independent, because returning to equilibrium magnetization also results in a loss of magnetization from the transverse plane (Deoni, 2010). Indeed, previous studies using the T1w/T2w showed a strong correlation with myelin volume fraction in human WM (Saccenti et al., 2020), oligodendrocyte-specific gene expression in humans (Patel et al., 2020), and with cortical myelin as compared to histological myeloarchitectural studies in both humans and non-human primates (Glasser & van Essen, 2011). However, others have found that the T1w/T2w correlated with MRI estimates of axonal diameter (Arshad et al., 2017), axonal density (Fukutomi et al., 2018), and iron content in healthy adults (Shams et al., 2019). Overall, evidence suggests that the T1w/T2w offers a promising measure of the functional aspects of WM microstructure (Uddin et al., 2019).

Because sensitivity or specificity of the T1w/T2w to myelin or axons is not known, our results need to be replicated along with more specific myelin imaging methods, such as myelin water imaging or quantitative magnetization transfer (Lee et al., 2020; MacKay & Laule, 2016).

### Limitations and future directions

Some methodological limitations warrant consideration. Because the simple division of the T1-WI by the T2-WI does not automatically cancel the different signal variations related to the different sensitivity profiles of the receiver coils of each image (Ganzetti et al., 2016), the variations in intensity of the MRI need to be corrected to obtain a standardized T1w/T2w contrast. Although the external calibration using non-brain tissues (i.e. eye and neck muscle) is considered the most robust (Ganzetti et al., 2016), we used the internal calibration proposed by Glasser and colleagues (Glasser & van Essen, 2011) due to insufficient brain coverage of our T2-W image to use the external hallmarks. The internal calibration rescales the image using the whole brain intensity distribution (Ganzetti et al., 2014; Glasser & van Essen, 2011). For this, we verified the resulting T1w/T2w pre and post calibration and our analysis yielded T1w/T2w histograms that had comparable intensity scales across subjects, in both the intervention and control groups. Still, our results need to be replicated using an external calibration approach.

Because we had insufficient brain coverage, we were not able to include other WM regions that have been associated with episodic memory, such as the WM tracts connecting the hippocampal formation (Burgess et al., 2002). Nevertheless, our study included subcortical connections of the hippocampus (e.g., fornix), and other direct projections (e.g., cingulate gyrus). Future research should include these relevant WM tracts to further characterize the relevant WM regions that can benefit from aerobic exercise interventions and can lead to increased episodic memory function.

Because the TBSS analysis used inherently focuses on normal appearing WM and the centers of the tracts, it may introduce bias by excluding some voxels affected by WM hyperintensities. We confirmed that the voxels sampled onto the WM skeleton from TBSS were derived from the same WM structures using the “deproject” function on FSL. Still, future studies should test different registration techniques to allow robust analyses in the entire WM.

## Conclusion

Our study provided the first evidence for the standardized T1w/T2w as a WM metric capable of detecting the effects of intervention-induced plasticity in the adult WM. Together, our findings suggest that the WM of the adult brain demonstrates plasticity in vulnerable regions and that these changes can be observed on a short-term scale. Still, more randomized controlled trials are needed to understand the heterogeneity of effects that aerobic exercise-based interventions have on the aging WM and cognitive function. Given that myelin-sensitive imaging MRI is not collected within the large studies on aging (e.g. ADNI(Jack et al., 2008), UK Biobank(Alfaro-Almagro et al., 2018), ENIGMA(Thompson et al., 2014), HCP (Sotiropoulos et al., 2013)) or randomized controlled trials (e.g. IGNITE(Erickson et al., 2019)), our findings suggest that T1w/T2w may offer an alternative and accessible metric of WM integrity. Our results encourage revisiting the existing datasets to further explore the potential of T1w/T2w to detect WM decline or plasticity.

## Data Availability

The data that support the findings of this study are available from the corresponding author, AZB, upon reasonable request.

## Acknowledgments

This work was supported by the National Institute on Aging at the National Institutes of Health (R37 AG025667), funding from Abbott Nutrition through the Center for Nutrition, Learning, and Memory at the University of Illinois (PIs Kramer and McAuley) and by the Translational Medicine Institute Translational Acceleration Program, Colorado State University (PI Burzynska). We would also like to thank Anya Knecht, Susan Houseworth, Nancy Dodge, Holly Tracy, and the Lifelong Brain and Cognition and Exercise Psychology Laboratory graduate students and staff for their help in participant recruitment and data collection, Michelle Hefner for helping with the image preprocessing and Vanessa Mendez for processing professionally all the illustrated figures.

## Disclosures

The authors declare no competing financial interests.

## Notes

### Competing Interest Statement

The authors have declared no competing interest.

### Clinical Trial

NCT01472744

