## Supplementary Material for "6-month aerobic walking training increases T1w/T2w signal in the white matter of healthy older adults"

**Supplementary Material 1**

*Flowchart diagram of sample selection*

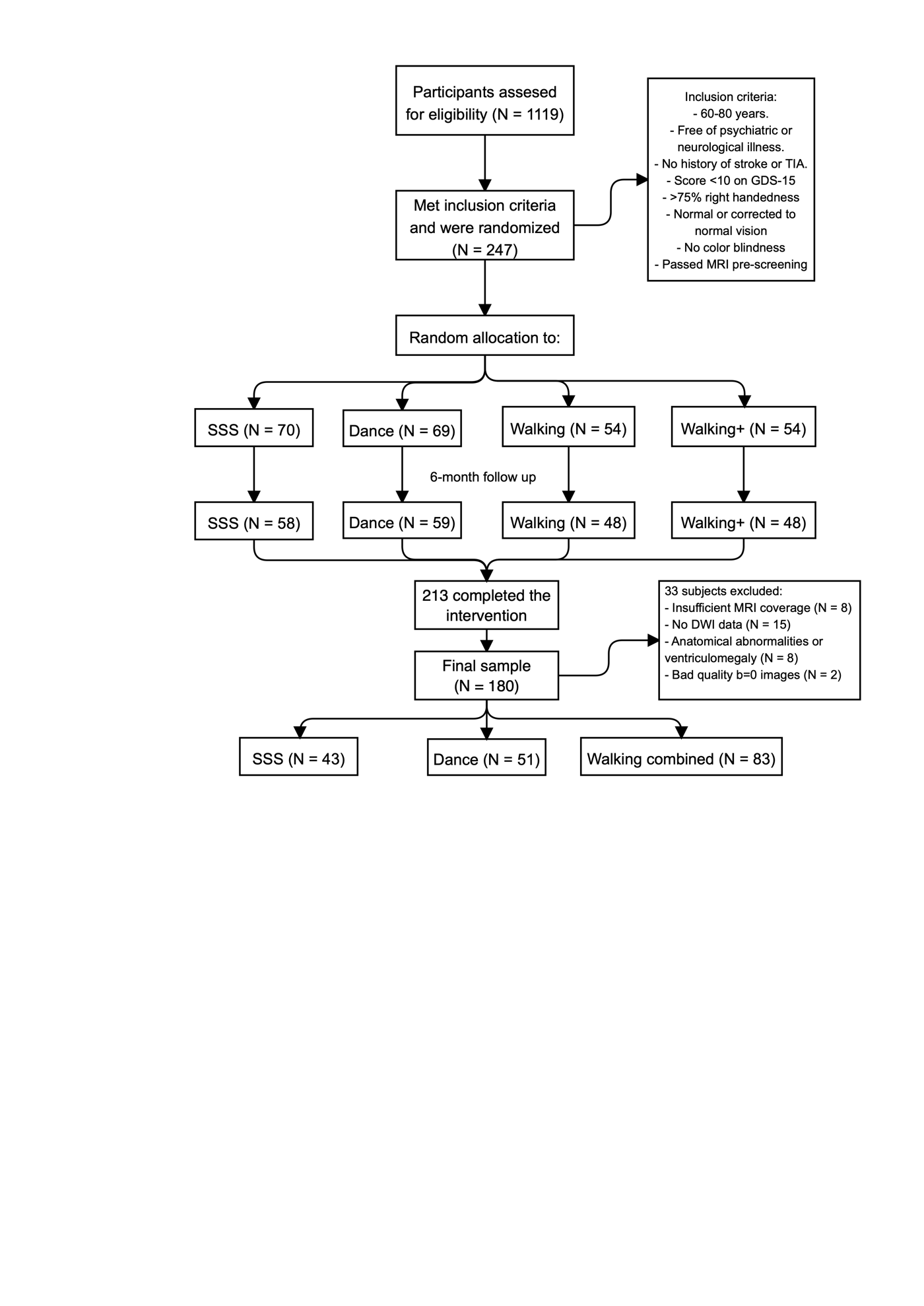

**Supplementary material 2**

*Time-by-group interactions in WM T1w/T2w*

|  | Walking + Dance vs. SSS | | | Walking vs. SSS | | | Dance vs. SSS | | |
| --- | --- | --- | --- | --- | --- | --- | --- | --- | --- |
| Region | *β* | SE | *p* | *β* | SE | *p* | *β* | SE | *p* |
| fMAJ | 0.03 | 0.03 | 0.26 | 0.03 | 0.02 | 0.34 | 0.02 | 0.03 | 0.4 |
| CST | 0.04 | 0.03 | 0.48 | 0.02 | 0.03 | 0.46 | 0.04 | 0.03 | 0.2 |
| UNC | 0.01 | 0.02 | 0.57 | 0.01 | 0.02 | 0.63 | 0.01 | 0.03 | 0.6 |
| EC | 0.02 | 0.03 | 0.49 | 0.02 | 0.03 | 0.53 | 0.02 | 0.03 | 0.6 |

| **Supplementary material 3**  *Baseline characteristics of the sample* | | | | |
| --- | --- | --- | --- | --- |
| Variables | SSS | Dance | Walking | *p* value |
|  | n=43 | n=51 | n=86 |  |
| **General characteristics** | |  |  |  |
| Age | 66.3±4.5 | 65.8±4.6 | 64.8±4.2 | 0.143 |
| Women, n (%) | 26 (65.0) | 37 (75.5) | 54 (67.5) | 0.508 |
| Education, yrs | 16.3±3.0 | 15.3±3.3 | 15.9±2.6 | 0.321 |
| MMSE | 28.5±1.4 | 28.4±1.5 | 28.5±1.4 | 0.879 |
| BMI | 30.4±6.1 | 30.5±5.9 | 30.4±4.9 | 0.993 |
| Systolic BP | 132.2±14.9 | 132.6±12.6 | 131.9±14.2 | 0.963 |
| Diastolic BP | 79.6±7.9 | 82.7±17.7 | 78.5±7.5 | 0.137 |
| CRF | 19.0±4.5 | 19.5±4.1 | 20.0±4.5 | 0.456 |
| **Cognition** |  |  |  |  |
| Memory | -0.05±0.9 | 0.01±0.7 | 0.02±0.8 | 0.900 |
| Speed | -0.21±0.8 | 0.05±0.8 | 0.07±0.8 | 0.178 |
| Executive Function | 0.03±0.6 | 0.06±0.6 | -0.06±0.6 | 0.488 |
| Vocabulary | 0.03±0.8 | 0.13±0.8 | -0.06±0.8 | 0.421 |
| **T1w/T2w levels** |  |  |  |  |
| Total | 1.37±0.1 | 1.40±0.1 | 1.39±0.1 | 0.125 |
| CC1 | 1.46±0.1 | 1.48±0.1 | 1.48±0.1 | 0.411 |
| CC2 | 1.33±0.2 | 1.34±0.2 | 1.37±0.2 | 0.183 |
| CC3 | 1.12±0.3 | 1.13±0.3 | 1.18±0.2 | 0.176 |
| CC4 | 1.03±0.3 | 1.05±0.3 | 1.08±0.3 | 0.494 |
| CC5 | 1.43±0.2 | 1.43±0.2 | 1.45±0.2 | 0.745 |
| prefrontal | 1.42±0.1 | 1.44±0.1 | 1.44±0.1 | 0.324 |
| fMAJ | 1.12±0.2 | 1.15±0.2 | 1.13±0.2 | 0.730 |
| fMIN | 1.40±0.1 | 1.42±0.1 | 1.41±0.1 | 0.381 |
| Cingulum | 1.38±0.1 | 1.41±0.1 | 1.41±0.1 | 0.129 |
| CST | 1.25±0.2 | 1.26±0.1 | 1.26±0.1 | 0.766 |
| SLF | 1.35±0.1 | 1.39±0.1 | 1.38±0.1 | 0.129 |
| FX | 0.81±0.1 | 0.81±0.1 | 0.83±0.1 | 0.642 |
| UNC | 1.22±0.2 | 1.29±0.2 | 1.24±0.2 | 0.882 |
| *Note.* MMSE= Mini-mental state examination, BMI= body mass index, BP=blood pressure, CRF=cardiorespiratory fitness. Quantitative data is presented as mean and standard deviation (±) and qualitative data as frequencies and percentages. *p* value of the comparison of the baseline information between SSS vs. dancing vs. walking. | | | | |

**Supplementary Material 4**

*Time-by-group interaction coefficients to compare Dance vs. Walking interventions*

|  | **Dance vs. Walking** | |
| --- | --- | --- |
| Region | Standardized β | *p* |
| Total | 0.02 | 0.844 |
| CC1 | -0.03 | 0.701 |
| CC2 | -0.01 | 0.996 |
| CC3 | -0.01 | 0.873 |
| CC4 | 0.02 | 0.598 |
| CC5 | -0.08 | 0.138 |
| Prefrontal | 0.01 | 0.867 |
| fMAJ | -0.01 | 0.722 |
| fMIN | -0.01 | 0.877 |
| Cingulum | -0.02 | 0.636 |
| CST | 0.02 | 0.485 |
| SLF | 0.02 | 0.728 |
| FX | -0.02 | 0.550 |
| UNC | -0.01 | 0.891 |
| EC | -0.01 | 0.975 |

**Supplementary material 5**

*Baseline correlation coefficients between T1w/T2w and cognition*

| Region | **Memory** | **Speed** | **Executive function** | **Vocabulary** |
| --- | --- | --- | --- | --- |
| Total | 0.09 | -0.01 | 0.05 | 0.13 |
| CC1 | -0.09 | 0.14 | 0.12 | -0.08 |
| CC2 | -0.09 | 0.14 | 0.05 | -0.19 |
| CC3 | -0.04 | 0.06 | -0.04 | -0.19 |
| CC4 | -0.06 | 0.08 | -0.02 | -0.16 |
| CC5 | -0.06 | 0.08 | 0.02 | -0.15 |
| Prefrontal | 0.15 | 0.02 | 0.10 | 0.17 |
| fMAJ | -0.08 | -0.03 | -0.04 | -0.15 |
| fMIN | 0.07 | 0.04 | 0.07 | 0.12 |
| Cingulum | 0.08 | 0.07 | 0.11 | 0.13 |
| CST | 0.15 | -0.08 | -0.02 | 0.10 |
| SLF | 0.18 | -0.03 | 0.06 | 0.21 |
| FX | -0.01 | 0.12 | 0.04 | 0.00 |
| UNC | 0.14 | -0.06 | 0.01 | 0.12 |
